# Challenges and benefits associated with educational materials: experiences of the Ghanaian nurse trainees

**DOI:** 10.1101/2024.06.30.24309743

**Authors:** James Agamah Adabre, Victoria Bubunyo Bam

## Abstract

**Background:** Educational Materials (EMs) are vital in teaching and learning activities and contribute significantly to the acquisition of knowledge and skills for an enhanced performance. The right educational resources help stimulate, reinforce and make it more impact and beneficial.

**AIM:** This study’s main goal is to identify the learning materials that are used regularly in nursing training schools and analyze the experiences of nurse trainees on their impact on the achievement of academic objectives. And to guide stakeholders in nursing education policy formulation.

**SETTING:** This study was conducted in four (4) Nursing and Midwifery Training institutions in Ghana.

**Methods:** A descriptive method was used for this study. An online survey was used to collect data from 47 nurse educators and 374 student nurses and analyzed using Stata version 16.0. The Kwame Nkrumah University of Science and Technology’s Committee on Human Research, Publication, and Ethics (CHRPE) was consulted for ethical approval.

**Results:** The common available EMs were handouts (80.1%), recommended textbooks (55.9%), and smartphones (54.6%). The EMs that were commonly used by students were the handout (58.6%), recommended textbooks (39.6%), and both internet information and PowerPoint slides (18.8%) each. Majority of the students (64.1%) reported that the commonly used EMs made good contributions towards their academic performance and 22.6% indicated that the used EMs made excellent contributions to their academic performance. Example e-books has OR of 0.43 and a p-value of 0.021.

**Conclusion:** Modern Ems such as reliable internet access, 3D animation videos should be made readily available in schools to greatly help to improve the standards of nursing education and contribute to quality healthcare workforce.

**Contributions:** Educational materials plays a vital role in health trainees’ education, increases nurses knowledge, and output on health care delivery.

## 1. Introduction & Background

Education being dynamic has evolved immensely over the years and similarly has nursing education evolved. Today, varieties of educational materials (EMs) are used worldwide. Some of these EMs include textbooks, handouts, pamphlets, smart-phones, tablet terminals, and personal computer (PC). According to Gusti et al, blended learning adds value to the teaching and learning experience, they added Students and/or faculty who are used to a traditional teaching and learning style may struggle in their own learning-teaching process (1), Mei & Lun, 2021). According to El-banna et al., 2017 prudent application of instructional pedagogy with abstract contents is recommended in accelerated programs. They also added that teachers should make available to students adequate resource and justification for their use (3).

In a changing healthcare context, there is an increased mandate to provide quality education that produces a knowledgeable and skillful workforce who delivers excellence and evidence-based care to clients. Nursing education plays a significant role in the production of nurses with effective competencies. Upgrading nursing and midwifery education are vital in improving workforce numbers, quality of health care, and the health systems as a whole (4). Not only that but for the fact that good educational system is a conditions necessary for the scientific development of a country (5). Nevertheless, the quest to determine the most appropriate EMs within nursing is still being sought by many nurse educators (6). Despite the variety and importance of EMs to academic performance of students, students still complain of being trained certain principles that seem to be non-concrete in nature without educational materials to support them when they are learning on their own. The results of a study commissioned by the Nursing and Midwifery Council of Ghana (NMC-Ghana) indicated that most libraries of nursing institutions are in deplorable state to support students’ academic performance effectively (7). Amankwaa and colleagues also assessed some variables that predict success in Nursing Licensure Examinations in Ghana. They found the following contributing factors for the difference in students’ academic performance; tutor factors and issues concerning the program may affects students’ academic performance. They suggested further studies should be done on the availability of academic resources to improve learning(8).

Technology is changing the structure of nursing education, with nurse educators’ knowledge and use of modern teaching practices in the classroom are much anticipated for nursing students encouragement, also students’ needs are essential to providing successful integration of technology in modern education (9). Therefore incorporating information and communication technology (ICT) into the classroom can make learning more engaging, motivating, and effective (10). It is therefore vital to seek approaches that make best use of the classes and transform the classroom into a platform for relations between teachers and students and critical thinking (11), Harerimana, 2019). According to Bvumbwe’s 2018 integrated review on nursing education challenges and solutions in Sub Saharan Africa indicated that most Sub Saharan Africa are facing similar problems in health education ranging from over stretched training institutions by high number of students, limited number of staff, inadequate infrastructure and resources, coupled with high demand on hospitals for clinical training (4). However solutions for the above in nursing education were found to be similar (4). Based on the foregoing, this study aimed to identify “commonly used educational materials and perception of nurse trainees in Ghana. Because notwithstanding the variety and importance of EMs to academic performance of students, it is observed that perception of nurse trainees on the use of educational materials have not been researched into in Ghana. Nursing education in Ghana, even though has improved, has challenges which increase the focused to the requirements of nursing education. The findings will serve as a guide for stakeholders in nursing education in policy formulation, improve on EMs or make changes where necessary. The students as individuals will also identify the common available learning materials that best suit their learning style which will help them achieve their academic goal.

## 2. METHODOLOGY

### 2.1 Study Design and Setting

This was a cross-sectional study conducted in four (4) Nursing and Midwifery Training institutions in the Bono and Ahafo Regions of Ghana. Each of the regions has four (4) Nursing and Midwifery Training Institutions making it a total of eight institutions. Two institutions from each region were used for this study.

### 2.2 Study Population

All diplomas in nursing students in nursing training institutions in the Bono and Ahafo Regions constituted the target population for this study.

### 2.3 Sampling and Sample Size Determination

Convenience and stratified sampling techniques were used to collect data for this study. A total of 194 second year and 180 third year students of the selected institutions were used for this study. The sample size for the survey was determined using the Yamane formula for sample size calculation (12). Using an estimated population of 373 second year and 324 third year students based on the institutions’ registry, a sample size of 374 respondents was estimated for the study. The possibility of making type I error was estimated at 5% with 95% confidence interval.

### 2.4 Study Instrument

A questionnaire was sent to the participating students, the questions were both close-ended and open-ended questions for respondents. A self-administered questionnaire comprising of 29 questions was used. Five (5) Likert scales were used in the questions to assess respondents’ views on educational materials by asking the extent they agree or disagree with a specific question or statement. The questionnaire consisted of four parts. The first section was demographic information (age, sex, marital status, etc.) and the second part was questions that assessed the availability of EMs. The third part was on the utilization of common educational materials and the fourth part was questions pertaining to the perception of student on the effects of educational materials on their academic performance.

### 2.5 Data Collection Procedure

The principal investigator (PI) submitted an online questionnaire (Google form) to respondents with a link through WhatsApp platforms to the respondents. Google form setting made responses anonymous. Respondents were informed that taking part in the study was voluntary and they could withdraw anytime.

### 2.6 Ethical considerations

Letters were written to the heads of the various nursing training institutions to seek permission for the study and approval was received before the study was conducted in the institutions. Ethical approval was sought from the Committee on Human Research, Publication and Ethics (CHRPE) Ref: CHRPE/AP/012/21 of the Kwame Nkrumah University of Science and Technology. Every ethical guiding principle for human participants in research (e.g. informed consent, confidentiality, privacy, no harm, voluntary participation etc.) were observed strictly in this research. Participant information leaflet that include a consent form was included in the questionnaire and participants answering the questionnaire meant they had consented.

### 2.7 Data Analysis

Stata version 16.0 was used for the analysis of the data. Descriptive statistics was performed, and frequencies and percentages were calculated for each item in the questionnaire. Factors associated with the utilization of educational materials, socio-demographic characteristic, and privately owned educational materials and their self-reported improved academic performance were determined using Logistic regression analysis. A confidence interval was computed at 95% and a p-value of 0.05 or less was considered as statistically significant.

### 2.8 Reliability & Validity

The face and content validity of the instruments of the questionnaire was assessed by an expert in the subject area. The questionnaire was also tested for reliability during the pretest to ensure that respondents understood the questions. The feedbacks from the pre-test of 15 students were incorporated in the questionnaires before the start of field work. Scrutiny of the pre-tested data was also done for accuracy, relevance, completeness, uniformity and consistency of the questions for the respondents. Cronbach alpha was used to determine the interrelatedness between the test questions. The reliability measurement for the questionnaire was high (Cronbach’s α = 0.96) for students.

## 3. Results

### 3.1 Socio-demographic characteristics of participants

Table 1 below gives the summary of the student respondents. Out of 374 expected respondents, three hundred and twenty-two 322 representing (86.1%) answered and submitted the questionnaire for analysis. One hundred and ninety-two (199) of the participants represented the second-year students whiles 123 represented the third-year students.

**Table 1.** Socio-demographic characteristics of students.

| **Variable** | **Frequency (n=322)** | **Percent (%)** |
| --- | --- | --- |
| **Sex** |  |  |
| **Male** | 77 | 23.9 |
| **Female** | 245 | 76.1 |
| **Age (years)** |  |  |
| **18 – 20** | 20 | 6.2 |
| **21-25** | 256 | 79.5 |
| **26-30** | 44 | 13.7 |
| **31-35** | 2 | 0.6 |
| **Year in school** |  |  |
| **2nd year** | 199 | 61.8 |
| **3<sup>rd</sup> year** | 123 | 38.2 |

### 3.2 Available educational materials

On EMs that the students have access to in school or owned privately are indicated in Table 2 below, 287(89.1%) students agreed to their school library being well resourced with materials to equip them study. These libraries are also opened for students use between 8 to 24 hours from Monday to Fridays. Internet connection on campus was a B plus as was confirmed by a total of 176 (54.8%) students. The percentage of internet speed however segregated as 11.1% reported difficulty in connecting, 40.7% reported slow connection. 44% confirmed that the connection was good and 3.7% reported high speed internet connectivity. Some Educational materials (EMs) are owned by the students these include: PC, Tablets, and Smart Phones being owned by the students as presented in table 2 below.

**Table 2:** Availability of educational resources.

| **Educational Resources** | **Frequency (n)** | **Percent (%)** |
| --- | --- | --- |
| **School's library is well resourced (n=321)** |  |  |
| **Disagree** | 31 | 9.7 |
| **Neither agree nor disagree** | 3 | 0.9 |
| **Agree** | 287 | 89.1 |
| **Internet connection in school (n=321)** |  |  |
| **No** | 145 | 45.2 |
| **Yes** | 176 | 54.8 |
| **Educational materials available in the school** |  |  |
| **Recommended textbook (n=322)** | 180 | 55.9 |
| **Personal computer (n=322)** | 69 | 21.4 |
| **School provided Laptop (n=322)** | 114 | 35.4 |
| **School provided Tablet (n=322)** | 24 | 7.5 |
| **Handout (n=322)** | 258 | 80.1 |
| **Audio CDs (n=322)** | 8 | 2.5 |
| **Video CDs (n=322)** | 5 | 1.6 |
| **Educational resources owned by students** |  |  |
| **Students own Laptop (n=322)** | 161 | 50.0 |
| **Tablet/Personal computer (n=322)** | 17 | 5.3 |
| **Smart phone (n=322)** | 176 | 54.6 |

### 3.3 Commonly used educational materials

A greater number of students 164 (56.0%) however would mostly use handouts from tutors and as low as 15 (5.1%) rarely uses it, 89(30.4%) frequently uses it and 25 (8.5%) least uses handouts from tutors. The second mostly used material is textbooks 60 (23.0), followed by information from the internet (58), and 4(1.8%) mostly only use audio CDs and as high as 163 (73.1%) least using it followed by video CDs with 155 (68.6%). 42 students least use PowerPoint, 26 frequently use it, 43 mostly use it, and 33 rarely use it. Soft copies of textbook usage was also assessed, 28 mostly use soft copies of textbooks, 43 frequently use it, 83 least use it, and 81 (34.5) rarely use it.

### 3.4 Utilization of the common available educational materials

Students were first asked to provide two books that were used for the core subjects they had done or were taking and indicate how frequently they used them. About 70% did not cite any textbook at all but rather wrote handout for all the subjects indicating that handouts were the educational materials used by majority of the students for studies. It was also found that certain books were used for more than one subject and majority of them ranges between 1-6 years and few being between 1-3 years old. Table 3 below shows the rate at which students use these common educational materials.

**Table 3.** Students’ utilization of commonly available educational resources.

| **Educational Resources** | **Not at all**<br>**n (%)** | **Minimally**<br>**used**<br>**n (%)** | **Moderately**<br>**used**<br>**n (%)** | **Often used**<br>**n (%)** |
| --- | --- | --- | --- | --- |
| **Information from the internet (n=290)** | 60 (20.7) | 96 (33.1) | 72 (24.8) | 62 (21.4) |
| **Recommended textbooks (n=284)** | 47 (16.6) | 76 (26.8) | 102 (35.9) | 59 (20.8) |
| **Handouts (n=306)** | 27 (8.8) | 19 (6.2) | 103 (33.7) | 157 (51.3) |
| **Soft copies of text books (n=270)** | 82 (30.4) | 108 (40.0) | 55 (20.4) | 25 (9.3) |
| **Visual aid (n=254)** | 109 (42.9) | 81 (31.9) | 36 (14.2) | 28 (11.0) |
| **Audio (n=251)** | 191 (76.1) | 35 (13.9) | 15 (6.0) | 10 (4.0) |
| **Personal computer (n=275)** | 76 (27.6) | 92 (33.4) | 67 (24.4) | 40 (14.6) |
| **Video CDS (n=251)** | 174 (69.3) | 48 (19.1) | 18 (7.2) | 11 (4.4) |

### 3.5 Reasons why students rarely use some EMs

On reasons why students rarely use some EMs, nine (9) variables were used for this exercise. They included audio, softcopy of textbooks, personal computer, video, handouts etc. As shown in table 4 below, Video (54.3%) and audio (57.6%) were hardly used because they did not have access to them. Students indicated that getting recommended textbooks was a waste of study time (14.2%) since tutors set questions from hands presented by the tutors in their respective intuitions. In relation to cost, handouts (51.5%), recommended textbooks (49%) and accessing internet information (45.8%) were considered as the three top sources that are expensive.

**Table 4.** Reasons students rarely use some EMs.

| **EDUCATIONAL RESOURCES** | **LITTLE KNOWLEDGE IN ITS USE (%)** | **EXPENSIVE (%)** | **WASTE OF STUDYING TIME (%)** | **DON'T HAVE ACCESS (%)** |
| --- | --- | --- | --- | --- |
| Audio | 24.2 | 12.3 | 5.9 | 57.6 |
| Soft copy of textbooks | 26.9 | 34.7 | 13.2 | 25.1 |
| Personal computers | 28.8 | 38.6 | 9.3 | 23.4 |
| Video | 21.9 | 17.1 | 6.7 | 54.3 |
| Information from internet | 28.4 | 45.8 | 8.5 | 17.4 |
| Recommended textbooks | 27.9 | 49 | 14.2 | 8.9 |
| Handouts | 28.5 | 51.5 | 12 | 8 |
| E-books | 21.5 | 31.1 | 9.1 | 38.4 |
| Visual aids | 26.2 | 23.8 | 11 | 39 |

### 3.6 Means of acquiring educational materials

Figure 1. Below gives a summary of the means by which students acquire educational materials for learning.

**Figure 1:**
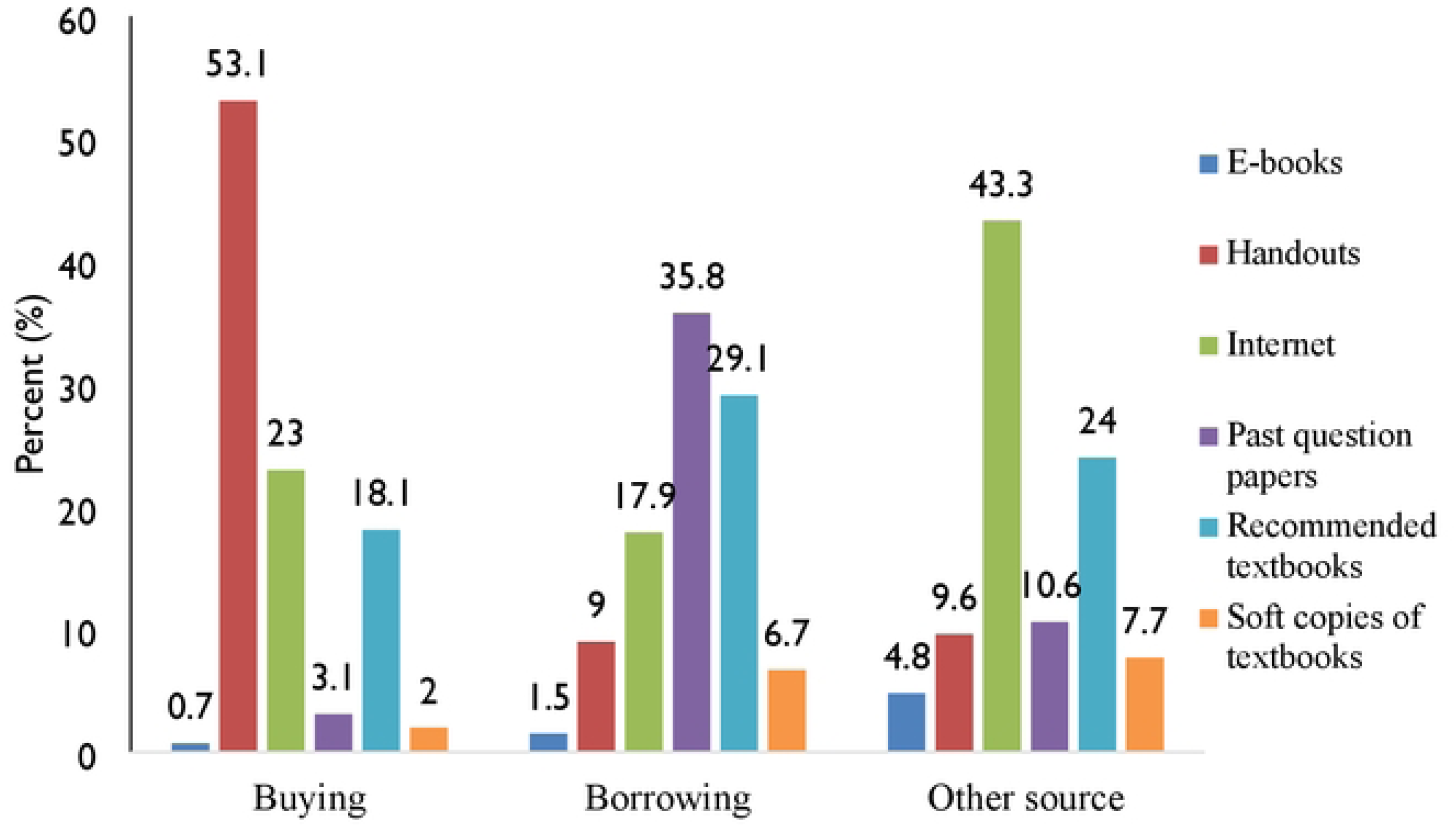
Means of acquiring learning materials.

### 3.7 Reasons for the use of type of the educational materials

Table 5 below, shows reasons for the use of type of the learning materials. It is indicated that 211(65.6%) students use the kind of learning material because of it is easy to understand the content, 139 (43.2%) students use their kinds because they are fast to read, and also easy to carry around, followed by how current the information is (50.3%). One hundred and six (39.1%) use what they believe tutors set their examination questions from and 120 (37.3%) use it for the reason that it is recommended by a tutor.

**Table 5:**
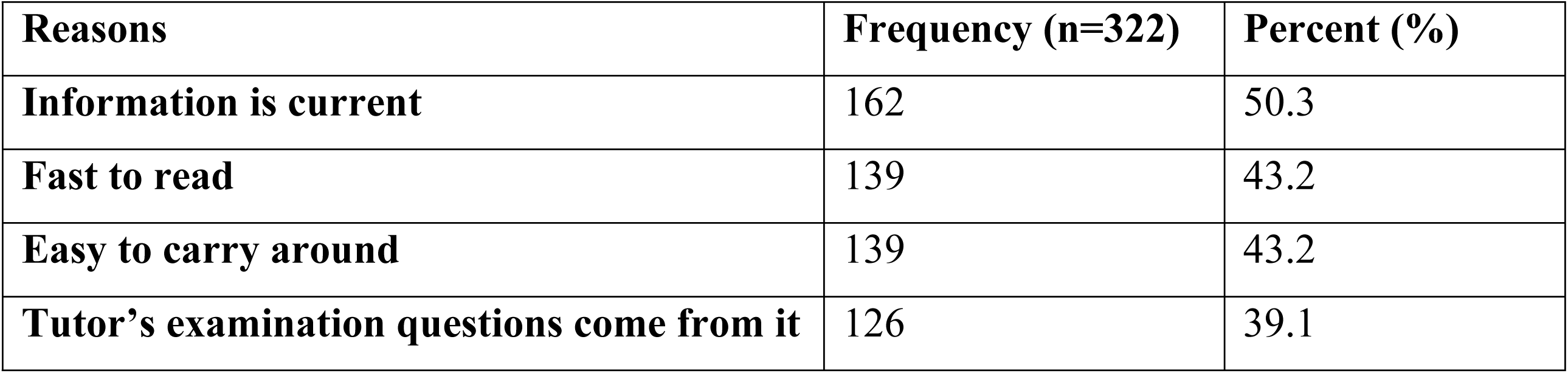
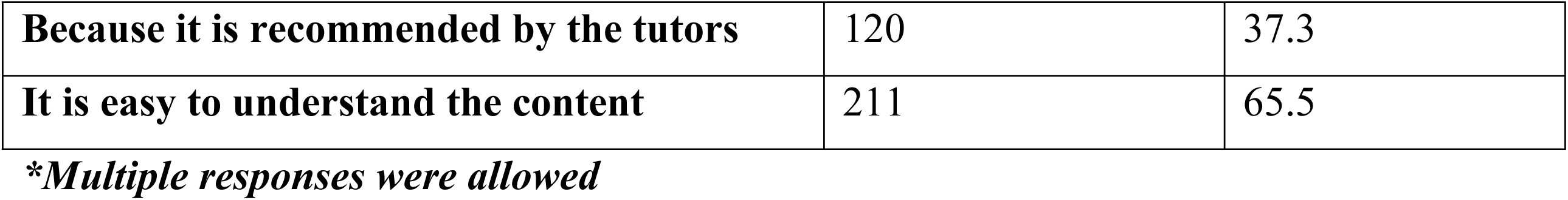
Reasons for the use of type of the educational materials.

### 3.8 Reasons student do not use some of the ems

On the reasons student do not use some of the EMs for their academic work. Majority of the students 168 (52.2%) indicated that slow internet access is a reason they don’t use internet as a source of information for their studies. A total of 155(48.1%) gave their reason as inability to afford, unavailability of the educational materials was cited by 114 (35.4%) students, 15.5% of the students also reported that examinations questions do not come from them, and the least (38 students) indicated that the information is outdated.

### 3.9 Challenges students encounter in accessing educational materials

On difficulties students encounter when accessing information for their course, 58.6% of the respondents cited poor or no internet services as the main challenge they encounter when accessing information for their courses. 33.2% cited financial constrains as a challenge they have in accessing information for their course, also lack of textbooks were cited by 26%, and 38.4% indicate that handouts which are their main source of information comes late in the semester was cited by 139 students as the difficulty they encounter as shown in on figure 2 below.

**Figure 2:**
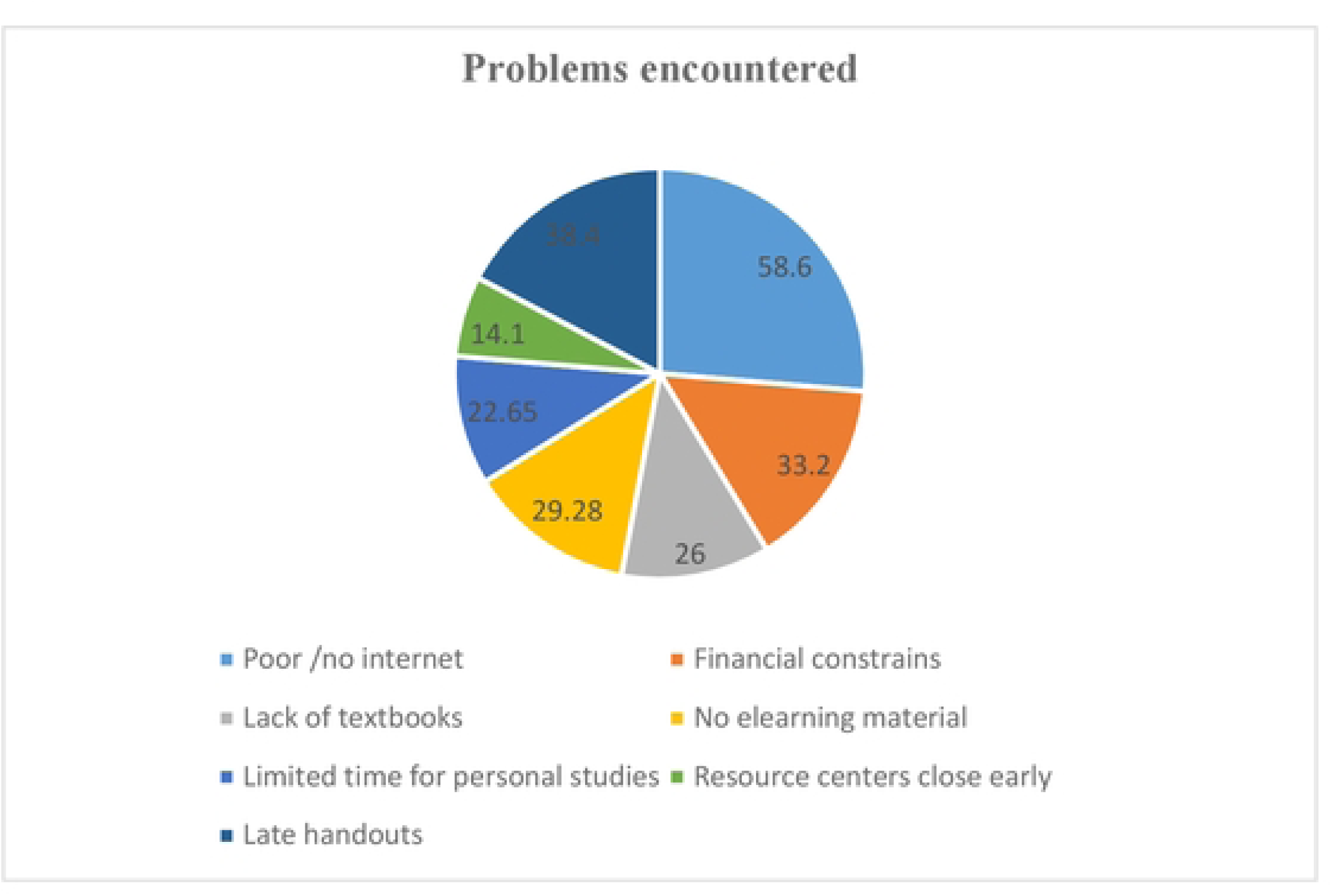
Challenges students encounter when accessing information for their courses.

### 3.6 Contributions of commonly used educational materials in general on students’ academic performance

Figure 3 below shows the contribution of educational materials to students’ academic performance. In this, four levels of contribution were used to perform this assessment and they included: not contributing, least contributing, good contribution and excellent contribution levels. Again educational material made impacts on students as 64.1% opted for Good contribution which was the highest suggested, 22.6% indicated that educational material makes excellent contribution to their academic performance.

**Figure 3.**
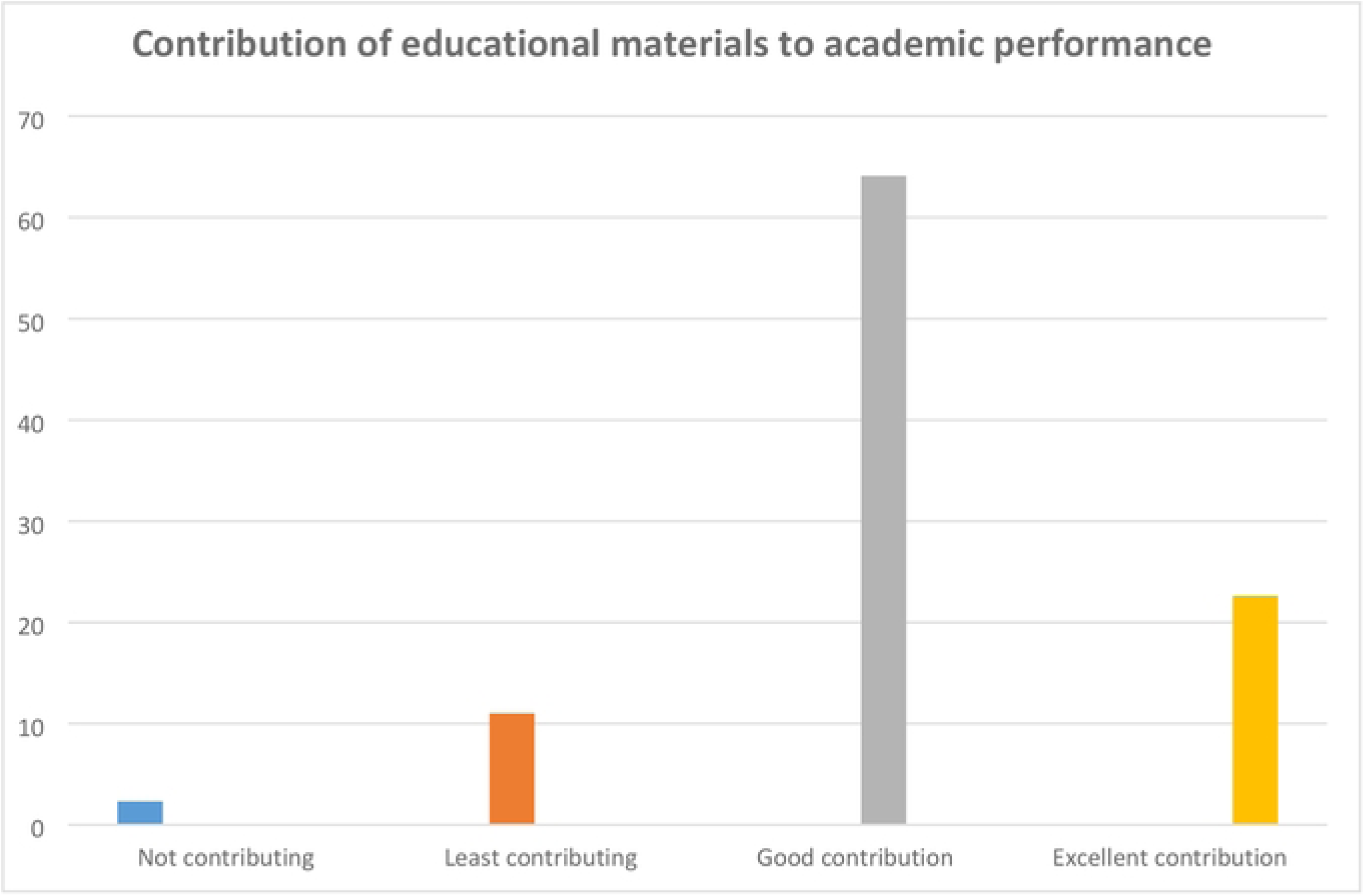
Contribution of EMs to academic performance.

### 3.7 Effect of commonly used educational materials on students’ academic performance

The study also assessed the nature of the effects that educational materials have on the students in their studies. Table 6 below shows that out of 273 respondents 142(52.0%) agree that EMs enhance their memory levels, 80(29.3%) strongly agree that educational materials enhances their memory level, 37(13.65) on the other hand disagree that EMs enhances their memory level. As to whether EMs improves students rate of accumulating knowledge, 16(5.8%) out of 274 strongly disagree whiles 88(32.1%) strongly agree. Out of 275, 96(34.9%) strongly agree that EMs meets the needs of students with different studying styles, 127(46.2%) agree, 42(15.3%) disagree, and 10(3.6%) strongly disagree.

**Table 6:** Students’ view on effect of commonly used educational materials on students’ academic performance.

| Effects of educational resources | Rating |  |  |  |  |
| --- | --- | --- | --- | --- | --- |
|  | Agree | Strongly agree | Disagree | Strongly disagree | Neither nor Disagree |
| It enhances the memory level of the students. | 142 (52.0%) | 80 (29.3%) | 37 (13.6%) | 14 (5.1%) | - |
| It facilitates the learning process. | 133(47.8%) | 103 (37.1%) | 32 (11.5%) | 10 (3.6%) | - |
| It improves students' rate of accumulation of knowledge. | 135(49.3%) | 88 (32.1%) | 35 (12.8%) | 16 (5.8%) | - |
| It Serves as tools used by the students that they cannot forget easily what they have learnt. | 137 (49.3%) | 93 (33.4%) | 36 (13.0%) | 12 (4.3%) | - |
| It brings about student concentration in the lesson | 132 (49.6%) | 89 (33.5%) | 31 (11.7%) | 14 (5.3%) | - |
| It permits the students self-evaluation. | 127 (47.7%) | 89 (33.5%) | 39 (14.7) | 11 (4.1%) | - |
| It meets the needs of students with different studying styles | 127 (46.2%) | 96 (34.9%) | 42 (15.3%) | 10 (3.6%) | - |

Logistic regression analysis was used in this analysis. At 95% confident interval, no significant relationship between utilization of educational materials and academic performance except e-books with p-value of 0.021. The students’ owning a tablets/personal computers reportedly had significant influence on academic performance (p<0.023).

## 4. Discussion

The aim of the study was to identify the commonly used educational materials and students’ perception about them on their academic performance in nursing training colleges in Ghana. Majority of the students considered that the most available and frequently used EMs made good contribution and excellent contribution to their academic performance. The most available or accessible among the materials are the print material (handouts and textbooks) followed by the electronic devices (smart phones and laptop).

Our finding had shown that even though internet connection on campus was rated high, the speed was inadequate. Gao and colleagues noted that a major problem faced by students throughout their usage of internet information was the discrepancy of contents from what was taught in class (13). Additionally, inadequate training, power supply and conducting assessments online are critical challenges in nursing education (14)

Aside libraries being opened for students and schools being connected to internet, students commonly use handouts, textbooks, smart phone, and laptops. Students use materials provided by the school or they own them. Handouts (print) are mostly used because it is easy to make changes and or corrections whiles effecting mistakes/typographical errors. Handouts are frequently used by nursing students more than recommended textbooks. On the contrary, according to (15), frequently used learning materials are visual displays. Our finding indicates that common educational materials not used at all or the least used are audio educational materials followed by video CDs. In the current study, students rarely use some of the educational materials because of inadequate access, high cost and the fact that they (students) have little knowledge in the use of certain educational materials. This is in line with the findings of Hendricks et al, (2017) that the high cost of educational materials is disturbing students’ academic performances, since they might not procure all the essential materials for their courses but would have to study without some of them (16).

Dahiya and Dahiya, ( 2015) found that students after only classroom teaching (chalk and talk) showed poor performance whereas most of the students possessed A+ to A (outstanding to very good) grades after implementing Classroom Seminar and Journal Club (CRSJC) model (17). This is in line with the current finding that educational materials contribute excellently in the nursing students’ academic performance. The current findings indicated that the collective reasons students do normally use textbooks frequently in their courses are to do assigned pre-readings, to help comprehend the material better than they could just from going to class alone, and to prepare for quizzes and exams. These findings are contrary to those of Hendricks et al. (2017) who indicated the reasons for the use of these EMs as: all the information one needs from class can be gotten, and much of the textbooks contain irrelevant material for exams (Hendricks, Reinsberg and Rieger, 2017).

To date, there has not been any single educational material has been found to achieve all curriculum requirements (18). It on this that Ismail 2021 indicate that Multimedia lecture presentations may increase students’ perception of important information and motivation for learning(19). Kabanga and colleagues also advocate for the use of diverse educational materials to enhance skill acquisition by nurses and midwives (20). Therefore it will be prudent to use multiple educational materials in nursing training colleges to get the desired results.

## 5. Limitations of the study

This study was impacted by the emergence of the COVID-19 pandemic which affected the opportunity for students to clarify any information and their responses, which could have explained some aspects of the data better. Other factors which influence academic performance were not explored by this study hence the results on academic performance cannot be wholly attributed to the perceptions and use of educational materials,

## 6. Recommendations

Educational materials in a form of tablets which contains the same learning materials should be made available for trainees across the country since every trainee in the country takes the same licensing exam, this will help make available authentic information for nursing students in the country. Additionally, students should use print materials simultaneously with other educational materials like electronic interactive. Also internet connectivity should be made compulsory in all nursing training institutions.

## 7. Conclusion

This study identified the commonly used educational materials, the extent of utilization of the educational materials, and assessed students’ perceptions on Educational Materials with regards to their academic performance in nursing training colleges.

It is time to institute measures that will make Ems readily available to teachers and students for evidence shows that there is the need for modern Ems in the training of nurses. This is for the fact that learning cannot be limited to knowledge of the teacher only.

## Authors’ contributions

Adabre James Agamah: Conception, design and writing of the manuscript.

Prof. (Mrs.) Victoria Bubunyo Bam: Critical reader, technical, administrative support, and reviewing

## Data Availability

All relevant data are within the manuscript and its Supporting Information files.

## Acknowledgement

We thank the management of all the schools for give us the permission to carry out this research, and the students for answering the questionnaires not forgetting of Augustine Lambonmung for always giving me assistance. May God bless you all.

